# Gender as a Social Determinant of Menstrual Health: A Mixed Methods Study Among Indian Adolescent Girls and Boys

**DOI:** 10.1101/2020.08.04.20167924

**Authors:** Mukta Gundi, Malavika Ambale Subramanyam

## Abstract

Gender bias in the patriarchal Indian society becomes evident in the form of worse sexual and reproductive health outcomes for girls than boys. While girls face menstrual taboos that affect their health, boys’ understanding of, and participation in, the menstruation discourse remains limited. We investigate how gender through its micro-interactional and macro-structural ways makes menstruation a gendered experience for adolescents; how various social determinants influence girls’ gendered menstruation experience across social domains; and whether the lived gendered experience of menstruation harms girls’ health.

Using a sequential mixed-methods design semi-structured interviews of 21 boys and girls each; 12 adult key-respondent interviews; and a cross-sectional survey of 1421 adolescents from urban, rural and tribal settings of Nashik district, India, were conducted. Applying social constructivist theory and gender analysis framework, we thematically analysed the qualitative data. Multivariable regression analysis of survey data yielded risk ratios.

Adolescents’ experience of menstruation was gendered. Fewer boys (versus girls) reported receiving information in schools [Incidence Rate Ratio (IRR) at 95% CI: 0.34 (0.24, 0.49)]. Girls’ gendered menstrual experiences varied across social domains and various socioeconomic backgrounds. Girls’ menstrual health was poorer among those with a lived experience of gendered menstruation [IRR: 0.22 (0.05, 0.90)]. Key respondents shared the need to engage boys in the menstruation discourse though apprehensive regarding its consequences. Gender bias along with other social factors negatively influence social construction regarding menstruation. Further, the discrimination is embodied by girls as poor health, thus perpetuating health inequalities across socioeconomic settings.

## Background

Indian girls and boys face several sexual and reproductive vulnerabilities that can harm their health during the transformative yet turbulent period of adolescence when their gender identities are still being shaped. While Indian girls showed a greater prevalence of genital infections and lower awareness regarding HIV/AIDS as compared to boys/young men (International Institute for Population Sciences (IIPS) and Macro international, 2007); a smaller proportion of boys than girls knew that a woman can get pregnant at first sex (International Institute for Population Sciences (IIPS) and Population Council, 2010).Additionally, our finding corroborates the NFHS-4 finding that girls’ constrained mobility and decision-making affects their healthcare seeking ability (International Institute for Population Sciences (IIPS) and Macro international, 2007; National Family Health Survey-4, 2017).

The resulting inequalities highlighted, frequently by sex-disaggregated datasets, are informative to policy-makers, yet incomplete unless gender itself is recognised as a social factor contributing to these inequalities. For instance, the observed differences could be wrongly interpreted as solely rooted in sex-linked biology (Dube, 1988; Gender and Health Group, 1999; Krieger, 2003; Morgan et al., 2016; Nowatzki & Grant, 2011). Moreover, to dig deeper into why certain social experiences, and opportunities, differ systematically for girls and boys, it is essential to understand the intersection of the processes driving gender inequalities with other socioeconomic inequalities (Hankivsky, 2012; Lorber & Moore, 2002). In the patriarchal Indian society, such interlinks create structural inequalities where girls and boys have unequal access to resources, and unequal norms hamper girls’ ability to question the gendered nature of their body and self (Dube, 1988; Thapan, 2001). Studies from Odisha show that girls face sanitation insecurity because physiological processes such as their menstruation and urination are governed by these intersecting inequalities (Caruso et al., 2017; Sahoo et al., 2015). For instance, socioeconomic inequalities were found to intersect with gender inequalities as women from lower socioeconomic strata could not find a private space to change their menstrual cloth or felt apprehensive as they faced drunken men while they walked to remote places to relieve themselves. Additionally, many Indian girls face menstrual taboos and strict norms within households and in the community; remain deprived of hygienic facilities at home and in public; and therefore, menstruation becomes a burden for girls (a Dasgupta & Sarkar, 2008; Kotecha et al., 2009; Narayan et al., 2001; P. Sharma et al., 2008; Singh, 2006). For instance, a study found that about 85% of school-going girls from West Bengal (n=160) faced menstruation-related restrictions (A. Dasgupta & Sarkar, 2008); about 75% of girls from Haryana (n=582) could not afford sanitary napkins ((V. Sharma et al., 2013); about one in four Indian girls miss at least a day in school during menstruation (van Eijk et al., 2016) and 43.46% older adolescent girls from Sikkim (n=1031) reported menstrual illness (Mishra & Mukhopadhyay, 2012).On the other hand, to our knowledge, except for a few qualitative studies, Indian boys are largely excluded from menstrual health research, furthering the understanding that menstruation is a “women’s topic “and missing avenues for including boys who not just play a role in health but in several facets of girls’ and women’s lives throughout the life-course (S.E. Mahon et al., 2015; Mason et al., 2013). A concern is that not recognizing menstruation as a gendered social experience that is potentially embodied by girls in the form of menstrual illnesses risks perpetuating gender inequalities in health.

Therefore, we investigate (Figure 1.1): (i) In what ways does gender determine adolescents’ menstruation-related knowledge, experiences and beliefs; (ii) whether girls’ social experience of menstruation is gendered and varies across social domains; (iii) whether a gendered experience of menstruation impacts girls’ menstruation-related beliefs and health.

**Figure 1.1:**
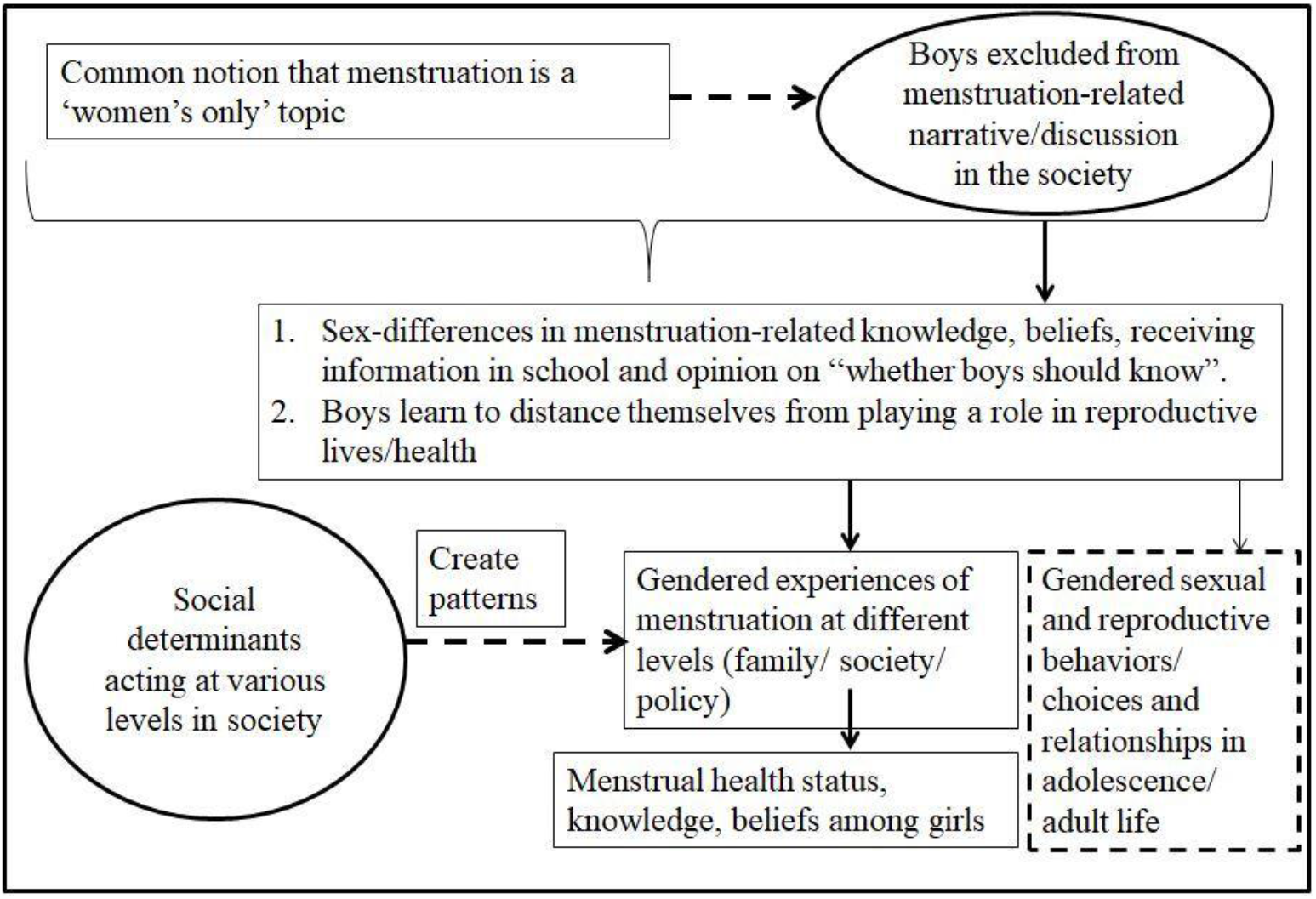
Conceptual Model for this chapter.

## Theoretical Frameworks

In addition to applying a broader lens of social construction of gender and the social determinants of health framework, we attempt at unravelling the minute ways in which adolescents learn and make sense of menstruation. To understand how menstruation is a gendered experience and how it impacts health, we used a gender analysis framework (Figure 1.2) that was put together by referring to: (i) the gender analysis toolkit for health systems (Jhpiego, 2016) and, (ii) the guidelines for analysing gender and health (Gender and Health Group, 1999).

**Figure 1.2:**
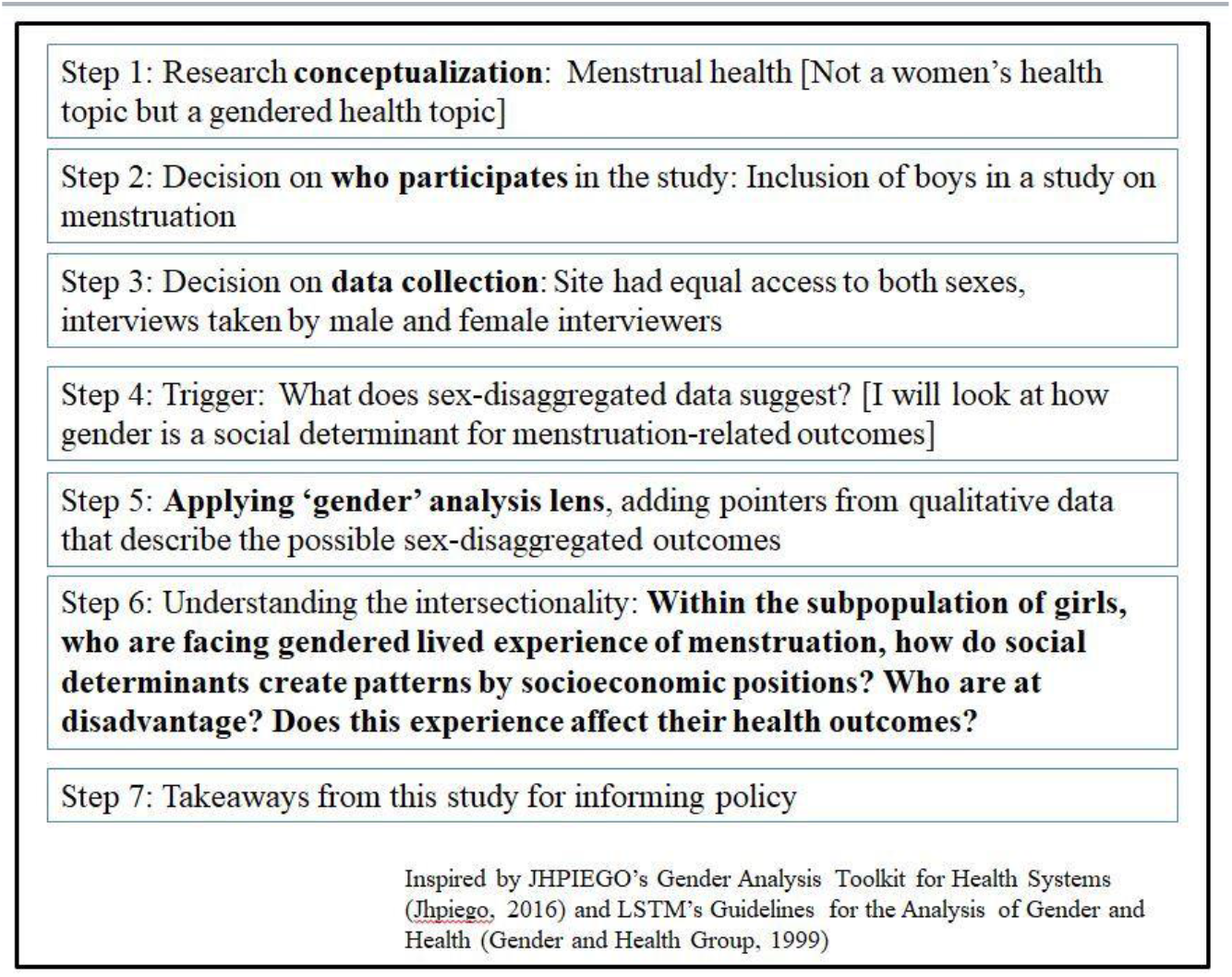
Gender Analysis Framework.

The theory of social construction of gender helped to understand the minute ways in which adolescent girls and boys, make sense of menstruation and its gendered nature (Fingerson, 2006; Tolman et al., 2003). To further study how it becomes a subject of concealment, we treated gender as a process by its ability to be produced and reproduced through everyday social engagements with its ability to order social life by creating subtle hierarchies (Fingerson, 2006; Lorber, 1994; Lorber & Moore, 2002; West & Zimmerman, 1987). This helped us theorize how gender through its micro-interactional and macro-institutional ways makes menstruation a discriminatory experience for girls and an exclusionary experience for boys (Ridgeway, 2009).

By using a gender analysis framework, we attempt to study how a gendered construction of menstruation operates and affects adolescent health (Nowatzki & Grant, 2011; Tolman et al., 2003). This framework helps examine gender power relations and its impact on menstruation-related health outcomes by questioning how gender-specific social roles, and society’s preferential inclusion and exclusion of a particular gender in the allocation of menstruation-related information and resources, influence adolescents’ social construction of menstruation in a step-by-step manner in the research process. We use gender-differences in menstruation-related outcomes as a trigger and question their unjustness (Gender and Health Group, 1999; Tolman et al., 2003).

## Methods Study site

Our research was carried out in the north-west part of Maharashtra state in India in Nashik district, with a contrasting characteristic of having a high Human Development Index (0.746) and some of the poorest inlands in the state (Jayachandran, 2014). The district has one of the largest tribal populations in the state with substantial socioeconomic heterogeneity (International Institute for Population Sciences (IIPS), 2014; International Institute for Population Sciences (IIPS) & ICF, 2017; Nashik District Collectorate, 2017).

## Study design

In this sequential mixed-methods study, we used a combination of an exploratory as well as explanatory approaches (Byrne, 2009). In the first phase, we conducted semi-structured interviews (adolescents aged 13-19 years and key-respondents) and focus group discussions (henceforth, FGDs) (separately for girls and boys). The emerged themes and observations from the qualitative data guided the survey questionnaire. A school-based cross-sectional survey using a sequential mixed methods design (Qual→ Quant→ qual), as described by Creswell et al. (2013), was carried out to capture the gender differences in menstruation-related outcomes, gendered experience and its impact on health among girls. A short post-survey qualitative phase was carried out to receive feedback on our survey findings. We integrated the quantitative and qualitative results to mix at the interpretation stage. This helped to bring forth the intricacies and patterns in the data as the complementary attributes of both the quantitative and qualitative (singular) methods were used (Onwuegbuzie & Johnson, 2006).

### Qualitative phase 1

Sampling of the 13-19 years old adolescent girls and boys was done using a mix of snowball and purposive sampling from three sub-district administrative divisions. Total 21 boys, 21 girls and 12 key-respondents (4 parents/ guardians, 3 teachers, 5 healthcare providers including a gynaecologist, an Ayurveda doctor, a homeopath, a nurse and a psychologist) were interviewed for about 25-45 minutes, nine FGDs lasting for 50-80 minutes were conducted in a familiar yet private place in schools, homes, or at workplaces (with the exception of one FGD that was conducted at a local non-governmental organization working on sanitation). Except for three boys’ interviews and one girls’ FGD who seemed nervous to record their responses, all other interviews were audio-taped.

In each interview and FGD, after a general introduction, participants were slowly directed towards the topic. Data were collected until data saturation was achieved. For this sensitive topic, we took care of the participants’ comfort by recruiting a male interviewer with a past experience with health-based grass-root level work in Maharashtra to talk to male participants. However, a female interviewer was present in addition to a male moderator for three interviews and three FGDs of boys. Through thick description for verbal as well as non-verbal responses we noted the perceived differences in boys’ responses for the presence/absence of the female researcher.

## Survey questionnaire formation

A field-test for the survey question-bank was carried out among 35 boys and girls. The questionnaire was modified according to the feedback received and was translated into Marathi; back-translated into English and was checked for content validity by experts.

## Survey

### Sampling and population

Initially, a list of schools stratified by setting (urban/rural/tribal) and type (government/ private aided/ private unaided) was prepared for participants’ selection from various socioeconomic settings (Goyal & Pandey, 2012). The sample size for the survey was calculated based on a maximum and the minimum expected prevalence for possible outcomes which was compared for adolescents’ subgroups using a two-sided test with 80% power. We required 1500 participants with 10% non-response, of which we could collect the data from 1491 participants. The analytical sample for the study was 1421 as the data of 70 premenarchal girls was excluded.

We could collect the survey data from 27 out of 35 approached schools through snowball sampling that covered about 2.5% of secondary schools in the district [28]. Students from different educational divisions with alternate roll numbers /alternate seats were chosen as participants. A research assistant (RA) and a school staff-member ensured smooth administration of the survey. In few cases, RA had to read out the questions in-person for participants with reading difficulty. More details regarding methodology could be found in another paper by (Gundi & Subramanyam, 2019).

#### Variables of Interest

The choice of the variables was guided by the findings from the pre-survey qualitative phase and the overall conceptual model for this study.

The outcome variables used were- knowledge (possesses/does not possess), and beliefs (favorable/unfavorable) regarding menstruation. Menstrual health status, a composite variable, was used as a binary outcome variable (no complaints regarding menstrual health/any complaint). The predictors used were (i) gender (boys/girls), **(ii)** received information in schools (yes/no) and (iii) comfort asking questions on menstruation to teachers (lack of complete comfort/ complete comfort) and (iv) the lived experience of gendered menstruation among girls, a composite variable operationalized as a binary variable (gendered experience/not) was used as a predictor. The gendered experience of menstruation (among girls) was captured using five separate variables depicting girls’ menstruation-related social experiences within households, schools, and communities.

##### Covariates

Age (early/late adolescents), caste [general (socially privileged caste-group)/other], and religion (Hindu/non-Hindu) and mother’s occupation (farmer, daily wage worker, employed, self-employed), and father’s occupation (farmer, daily wage worker, employed/self-employed, other) were used as covariates. The details regarding variables are found in Appendix 1.

Five girls (of which, 2 were married) and three key respondents (a female tribal-village head, a sex education expert and a social worker) were interviewed for post-survey qualitative phase, to seek their feedback on our study findings. Despite efforts, we were unable to recruit school dropout boys due to lack of parental consent.

## Ethical considerations

Ethical approval was obtained from the IIT Gandhinagar Institutional Ethics Committee prior to the study. School authorities were approached in advance to discuss survey objectives and detailed procedure. Prior verbal consent was obtained from parents/guardians by the school representatives, as suggested by the school authorities. After obtaining the consent, we were invited by the school authorities to collect the data. Participants were told the aim and the voluntary nature of participation. Verbal consent from students’ parents/guardians was requested to be acquired by the school representatives. Adolescents’ verbal consent/ assent (depending upon their age) was sought prior to their participation in the qualitative interviews/FGDs or survey. They could choose either English or Marathi survey form.

They were informed that they could ask any queries at any point and that they can ask us to stop the audio taping if they wished to speak off-the-record. They were also told that they could withdraw from the study at any point without any consequences. The anonymity of the survey and the confidentiality of the data was maintained throughout the study. More details are found in other paper (Gundi & Subramanyam, 2019).

## Analysis

### Qualitative data analysis

After we transcribed and translated the recorded data into English, we got the translation verified by an expert to ensure that the translation did not lose nuanced meanings. The data was then analyzed using thematic analysis (Braun et al., 2019). After a thorough reading of the transcripts, through open coding we identified meaningful statements or words with similar findings to create labels. Then through axial coding, any association between labels was identified (Terry et al., 2017). Later, we converted the selected codes into shorter codes and created the categories with shared meanings. We also ensured data triangulation by including member (participant) rechecking and peer checking. Additionally, a deep engagement with the participants and key participants’ interviews further strengthened data triangulation (Terry et al., 2017; Willig & Rogers, 2017).

### Quantitative data analysis

The data were cleaned, labelled and the descriptive statistics of the main variables were calculated. A generalized linear model with a logarithmic link function and binomial distribution for the residual were fit as a form of log binomial regression models that produced prevalence ratios for the binary dependent variables at an alpha value of 0.05. We accounted for school-level clustering by using robust standard errors in the regression analysis. In case the models did not converge, Poisson regression models with survey commands were used to model the incidence rate ratios (equivalent to prevalence ratios in this study). The unadjusted model (model 1) was adjusted for age, setting, gender (wherever they were significant) in model 2. Model 3, following our conceptual model, included parental socioeconomic status and school type. The findings were tested for the exclusion of non-significant predictors as a part of sensitivity analysis. STATA version 12 was used for data analysis (StataCorp, 2012).

## Results

### Qualitative phase 1

#### Gendered Experience of Menstruation in Different Domains of Life

Girls’ gendered social experience of menstruation cut across different social domains such as households, schools, and neighborhoods. Most girls faced severe restrictions within households than in schools/ communities as they were expected to hide their menstrual status and absorbent from men in the family.

Several menstruating girls across different settings and socioeconomic backgrounds were either prohibited from or felt awkward while interacting with men. Although restrictions were reportedly becoming less rigid as compared to earlier generations, a few girls expressed an inability to overrule or question these gendered norms.

> “I am not allowed to offer any food to men in the family and I am required to sit away during meal-time “
>
> “Why do you think you are required to follow this?
>
> “…umm.. (pause)… because of having periods…“
>
> “I understand, but why do you have to follow this when you have your periods?
>
> “..I am not really sure…“
>
> “Do you think you can ask this question to your mother?
>
> ““No…I don’t think I can… (smiles)“
>
> - Komal (14 years, urban setting)

However, this largely pervasive gendered experience was not found to be analogous in all domains. For instance, although embarrassment while buying sanitary pads from a male shopkeeper was narrated by a few girls from wealthy urban households, they did not report hiding their menstruation from men in the family. Conversely, a girl from an urban area who dried her absorbent cloth at home by hiding it under a bigger cloth was comfortable attending a sex-education workshop at her mixed-gender school.

#### Patterns in Girls’ Gendered Experiences of Menstruation

Girls’ gendered experience of menstruation was influenced by their socioeconomic setting and school-type. It was common for girls from non-urban areas to experience teasing from boys for menstruation. Girls from both urban and non-urban settings without private toilet facility seemed to accept that menstruation was not a private matter. Several girls from rural setting were not allowed to visit or touch anyone outside to respect commonly followed social norms. A few educated mothers too gave gender-specific instructions to their daughters. However, most girls from private unaided schools and wealthy families felt comfortable discussing menstruation in mixed-gender classrooms and in front of their fathers. Except two fathers from wealthy urban families, no father played an active role in menstruation.

> “..My dad told me about menstruation before my periods started, when I was in 6^th^ or 7^th^ standard I guess. He gave me a book to read and said, “Let’s discuss if you have any questions. “I read the book and then discussed many things that I did not understand.. He also took us for dinner when I got my first periods.. (Smiles)“
>
> - 15 years-old girl, urban setting

Our key respondent interviews showed that parents across varied socioeconomic backgrounds (except for two highly educated mothers) communicated gender-specific expectations to their menstruating daughters.

### Boys’ Role in Menstruation

An apprehension at the possibility of being teased by boys was palpable among most of the girls who opposed the idea of boys being informed about menstruation.

> “Do you think boys should be given detailed information about menstruation?”
>
> “No… not at all..”
>
> “No” (loud and a firm response)
>
> “No… I mean it is both good and bad at the same time..”
>
> “Why do you say that?”
>
> “Good thing is that they will know about it as it is important for future.. as they grow as adults.. but they will also laugh at us, tease us if they know more details”.
>
> “I think if they are told what menstruation is, they should also be told that it is not good to tease about it”.
>
> - Focus group discussion (Girls, Urban setting)

However, a few girls (mostly urban) stressed that boys should receive the information from appropriate sources as exploring wrong sources could misguide them. Questions about boys’ perceived role in menstruation yielded a mixed response from boys. Many boys expressed their role as altruistic “to help and behave when girls are in pain”, while only a couple of boys from an urban setting, who attended sex-education workshops in schools, argued that learning about menstruation should be natural so that boys engage in healthy sexual relationships. One of the boys (14 years old) said, “Right now we are in a protected environment. When we will enter college, there is a possibility that if we don’t know about these things, we may engage in risky behaviors that can harm girls.”

Although the adolescent psychologist supported this, most parents opposed including boys in menstrual discussion until they turned sixteen. Parents from disadvantaged backgrounds were unsure of any meaningful roles that boys and men could play even if they knew about menstruation.

## Quantitative phase

Below is a gender distribution of adolescents with adequate knowledge, favorable beliefs, receiving information in school, and opining favourably to ‘whether boys should know’ (Table 1.1).

**Table 1.1:** Gender distribution of adolescents with adequate and accurate knowledge, favorable beliefs, receipt of information in schools and favorable opinion regarding whether boys should know about menstruation (n= 744 boys and 677 girls)

|  | Possess adequate and accurate knowledge |  | Total | Possess favorable beliefs |  | Total | Received information in schools |  | Total | Whether boys should know |  | Total |
| --- | --- | --- | --- | --- | --- | --- | --- | --- | --- | --- | --- | --- |
|  | Girls | Boys | Total | Girls | Boys | Total | Girls | Boys | Total | Girls | Boys | Total |
| **Setting** |  |  |  |  |  |  |  |  |  |  |  |  |
| Rural | 31 | 10 | 41 | 15 | 25 | 40 | 73 | 29 | 102 | 34 | 48 | 82 |
| % | 75.6 | 24.3 | 100 | 37.5 | 62.5 | 100 | 71.6 | 28.4 | 100 | 41.5 | 58.5 | 100 |
| Tribal | 75 | 22 | 97 | 21 | 35 | 56 | 156 | 53 | 209 | 67 | 108 | 175 |
| % | 77.3 | 22.7 | 100 | 37.5 | 62.5 | 100 | 74.6 | 25.4 | 100 | 38.3 | 61.7 | 100 |
| Urban | 127 | 56 | 183 | 44 | 95 | 139 | 297 | 120 | 417 | 140 | 187 | 327 |
| % | 69.4 | 30.6 | 100 | 31.6 | 68.4 | 100 | 71.2 | 28.8 | 100 | 42.8 | 57.2 | 100 |
| **Wealth tertile** |  |  |  |  |  |  |  |  |  |  |  |  |
| Poorest | 72 | 23 | 95 | 20 | 39 | 59 | 141 | 60 | 201 | 64 | 104 | 168 |
| % | 75.8 | 24.2 | 183 | 33.9 | 66.1 | 100 | 70.2 | 29.9 | 100 | 38.1 | 61.9 | 100 |
| Middle | 64 | 16 | 80 | 26 | 39 | 65 | 158 | 40 | 198 | 64 | 94 | 158 |
| % | 80 | 20 | 183 | 40 | 60 | 100 | 79.8 | 20.2 | 100 | 40.5 | 59.5 | 100 |
| Richest | 83 | 40 | 123 | 27 | 66 | 93 | 176 | 85 | 261 | 92 | 111 | 203 |
| % | 67.5 | 32.5 | 183 | 29 | 71 | 100 | 67.4 | 32.6 | 100 | 45.3 | 54.7 | 100 |
| **Mother's Education** |  |  |  |  |  |  |  |  |  |  |  |  |
| No/some elementary school | 88 | 27 | 115 | 33 | 40 | 73 | 182 | 66 | 248 | 88 | 117 | 205 |
| % | 76.5 | 23.5 | 100 | 45.2 | 54.8 | 100 | 73.4 | 26.6 | 100 | 42.9 | 57.1 | 100 |
| Middle school | 66 | 26 | 92 | 22 | 49 | 71 | 165 | 53 | 218 | 60 | 112 | 172 |
| % | 71.7 | 28.3 | 100 | 31 | 69 | 100 | 75.7 | 24.3 | 100 | 34.9 | 65.1 | 100 |
| High-school/Some college | 30 | 11 | 41 | 12 | 27 | 39 | 85 | 31 | 116 | 35 | 51 | 86 |
| % | 73.2 | 26.8 | 100 | 30.8 | 69.2 | 100 | 73.3 | 26.7 | 100 | 40.7 | 59.3 | 100 |
| College/more | 47 | 24 | 71 | 11 | 36 | 47 | 83 | 48 | 131 | 53 | 55 | 108 |
| % | 66.2 | 33.8 | 100 | 23.4 | 76.6 | 100 | 63.4 | 36.6 | 100 | 49.1 | 50.9 | 100 |
| **Father's Education** |  |  |  |  |  |  |  |  |  |  |  |  |
| No/some elementary school | 59 | 19 | 78 | 19 | 27 | 46 | 52 | 119 | 171 | 55 | 68 | 123 |
| % | 75.6 | 24.4 | 100 | 41.3 | 58.7 | 100 | 30.4 | 69.6 | 100 | 44.7 | 55.3 | 100 |
| Middle school | 69 | 25 | 94 | 26 | 43 | 69 | 155 | 53 | 208 | 69 | 109 | 178 |
| % | 73.4 | 26.6 | 100 | 37.7 | 62.3 | 100 | 74.5 | 25.5 | 100 | 38.8 | 61.2 | 100 |
| Highschool/Some college | 36 | 13 | 49 | 9 | 35 | 44 | 107 | 34 | 141 | 31 | 73 | 104 |
| % | 73.5 | 26.5 | 100 | 20.5 | 79.6 | 100 | 75.9 | 24.1 | 100 | 29.8 | 70.2 | 100 |
| College/more | 63 | 31 | 94 | 20 | 47 | 67 | 121 | 65 | 186 | 75 | 84 | 159 |
| % | 67 | 33 | 100 | 29.9 | 70.2 | 100 | 65.1 | 34.9 | 100 | 47.2 | 52.8 | 100 |
| **Age** |  |  |  |  |  |  |  |  |  |  |  |  |
| Young adolescents | 93 | 21 | 114 | 40 | 35 | 75 | 221 | 45 | 266 | 93 | 95 | 188 |
| % | 81.6 | 18.4 | 100 | 53.3 | 46.7 | 100 | 83.1 | 16.9 | 100 | 49.5 | 50.5 | 100 |
| Older adolescents | 136 | 66 | 202 | 40 | 116 | 156 | 294 | 152 | 446 | 144 | 242 | 386 |
| % | 67.3 | 32.7 | 100 | 25.6 | 74.4 | 100 | 65.9 | 34.1 | 100 | 37.3 | 62.7 | 100 |
| **Type of school** |  |  |  |  |  |  |  |  |  |  |  |  |
| Government | 71 | 15 | 86 | 15 | 31 | 46 | 162 | 44 | 206 | 53 | 91 | 144 |
| % | 82.6 | 17.4 | 100 | 32.6 | 67.4 | 100 | 78.6 | 21.4 | 100 | 36.8 | 63.2 | 100 |
| Private aided | 113 | 40 | 153 | 56 | 74 | 130 | 162 | 44 | 206 | 137 | 174 | 311 |
| % | 73.9 | 26.1 | 100 | 43.1 | 56.9 | 100 | 78.6 | 21.4 | 100 | 44.1 | 55.9 | 100 |
| Private unaided | 49 | 32 | 81 | 9 | 49 | 58 | 271 | 87 | 358 | 51 | 77 | 128 |
| % | 60.5 | 39.5 | 100 | 15.5 | 84.5 | 100 | 75.7 | 24.3 | 100 | 39.8 | 60.2 | 100 |

Missing data: Age (14), information received from school (12), comfort with teacher (26), mother's education (17), father's education (19), school type (1), wealth tertile (79), attitudes (28), whether boys should know (18), "women are dirty" (39), "women are impure" (14), "menstruation is a curse" (16), agreement with religion restrictions (11), agreement with cooking restrictions (11), agreement with women sitting away (13), meaning of menstruation (27), physiological process of menstruation (18), when does menstruation begin (8)

Below are gender differences - for possessing adequate and accurate knowledge (n=677 girls and 703 boys), favorable beliefs (n=574 girls and 684 boys), receiving information on menstruation in school (n=673 girls and 732 boys) and opining favourably to ‘whether boys should know about menstruation’ (n=660 girls and 727 boys) (Figure 1.3)

**Figure 1.3:**
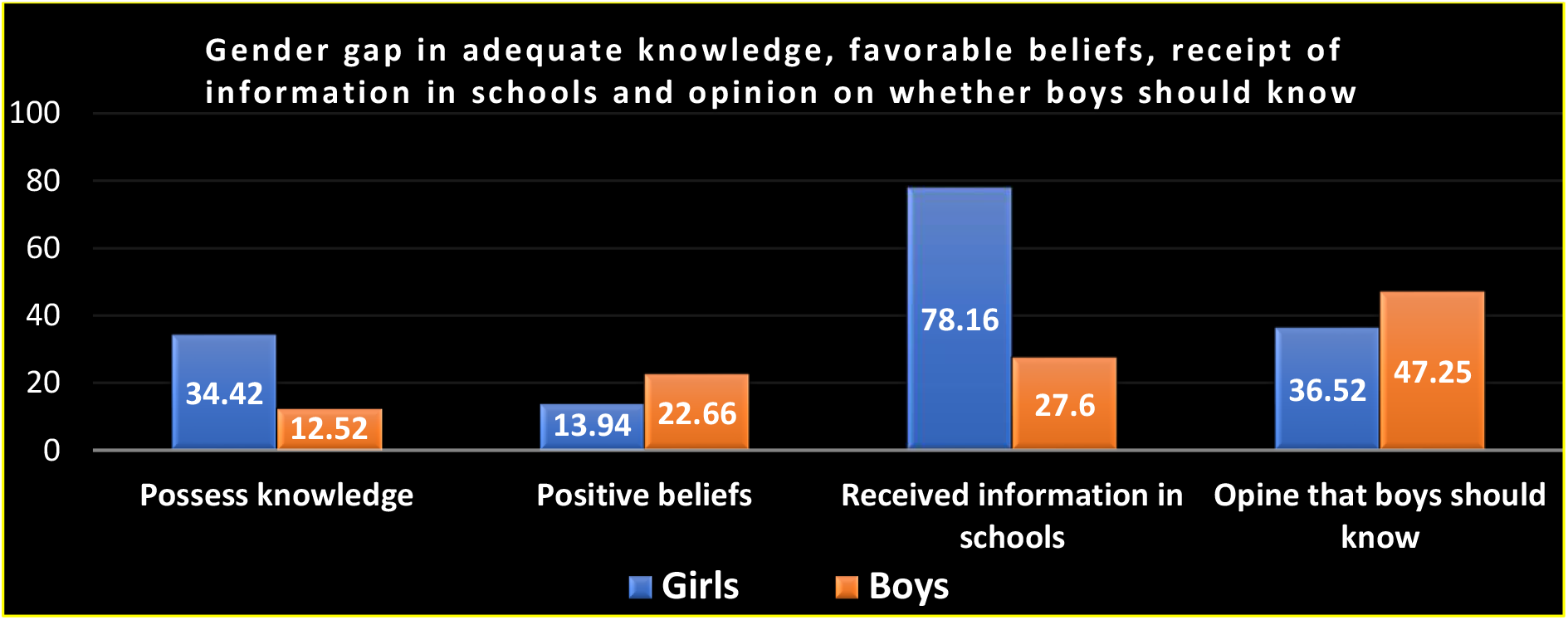
Gender gap in menstruation-related outcomes.

Notably, about 95% of the girls (n=443) reported at least one gendered social experience of menstruation. (Figure 1.4)

**Figure 1.4:**
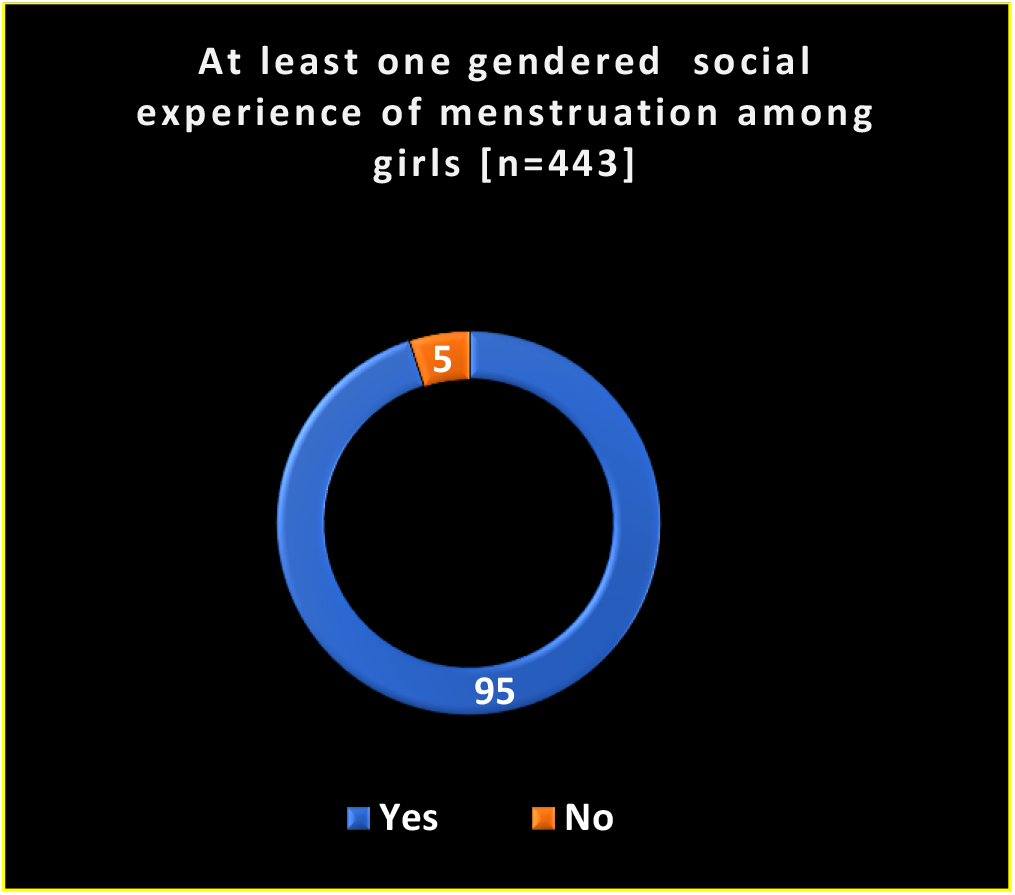
Percentage of girls reporting at least one gendered social experience of menstruation.

***Gender as a social determinant of menstruation-related outcomes among girls and boys (Table 1.2):***

**Table 1.2:** Incidence Rate Ratios (95% confidence intervals) of the association of gender with menstruation-related outcomes (n= 744 boys + 677 girls)

|  |  |  |  |  |  |  |  |  |  |  |  |  |
| --- | --- | --- | --- | --- | --- | --- | --- | --- | --- | --- | --- | --- |
|  | Knowledge (1=possess) |  |  | Positive beliefs (1=possess) |  |  | Received information in schools (1=received) |  |  | Whether boys should know (1=yes) |  |  |
|  | Model 1 | Model 2 | Model 3 | Model 1 | Model 2 | Model 3 | Model 1 | Model 2 | Model 3 | Model 1 | Model 2 | Model 3 |
| Gender |  |  |  |  |  |  |  |  |  |  |  |  |
| Girls | 1 | 1 | 1 | 1 | 1 | 1 | 1 | 1 | 1 | 1 | 1 | 1 |
| Boys | 0.36 (0.26, 0.50)*** | 0.35 (0.25, 0.49)*** | 0.34 (0.24, 0.47)*** | 1.62 (1.01, 2.59)* | 1.52 (0.95, 2.45) | 1.63 (0.99, 2.68) | 0.35 (0.24, 0.50)*** | 0.34 (0.24, 0.48)*** | 0.34 (0.24, 0.49)*** | 1.29 (1.05, 1.59)* | 1.26 (1.01, 1.57)* | 1.25 (1.00, 1.58)* |
| Notes: 1) Statistically significant p values indicated as *<0.05, **0.01, ***<0.001 2) Model 1: Unadjusted; Model 2: Age, setting adjusted; Model 3: Fully adjusted for age, setting, type, parental education and wealth. |  |  |  |  |  |  |  |  |  |  |  |  |

In fully adjusted models, fewer boys than girls possessed knowledge [IRR at 95% CI: 0.34 (0.24, 0.47)]. Although fewer boys than girls had received information in school [IRR at 95% CI: 0.34 (0.24, 0.49)], more boys (than girls) opined that boys should know about menstruation [IRR at 95% CI: 1.25 (1.00, 1.58)].

***Influence of gendered experience of menstruation on health-related outcomes (Table 1.3):***

**Table 1.3:** Incidence Rate Ratios (95% confidence intervals) of the association of the gendered experience of menstruation as a lived experience with girls’ health outcomes (n= 677 girls)

|  |  |  |  |  |  |  |  |  |  |
| --- | --- | --- | --- | --- | --- | --- | --- | --- | --- |
|  | Knowledge (1=possess) |  |  | Positive beliefs (1=possess) |  |  | Menstrual health status [1=no symptoms] |  |  |
|  | Model 1 | Model 2 | Model 3 | Model 1 | Model 2 | Model 3 | Model 1 | Model 2 | Model 3 |
| Gendered experience |  |  |  |  |  |  |  |  |  |
| No | 1 | 1 | 1 | 1 | 1 | 1 | 1 | 1 | 1 |
| Yes | 0.67 (0.46, 0.99)* | 0.67 (0.42, 1.05) | 0.68 (0.40, 1.14) | 0.39 (0.12, 1.24) | 0.40 (0.11, 1.43) | 0.39 (0.16, 0.93)* | 0.34 (0.11, 0.98)* | 0.33 (0.09, 1.13) | 0.22 (0.05, 0.90)* |
| Notes: 1) Statistically significant p values indicated as *<0.05, **0.01, ***<0.001; Missing data: Menstrual health status (46) |  |  |  |  |  |  |  |  |  |

A gendered experience of menstruation versus a lack of it was negatively associated with positive beliefs [IRR: 0.39 (0.16, 0.93)] and with good menstrual health status [IRR: 0.22 (0.05, 0.90)], in fully adjusted models.

## Qualitative Phase 2

Key respondents agreed with our study findings and suggested looking deeper into the social patterning of gender inequalities and its influences on menstruation-related experiences. Our interviews underscored the scope of capturing more shades of girls’ gendered experience of menstruation to understand how it impacts their lives. For instance, a 17-year-old unmarried girl from the tribal setting shared that

> “Our male teacher locks the toilet of the school and keeps the key with him.. he denies us access to toilet saying that we make it dirty…”. A similar experience was shared by another 19-year-old girl from tribal setting who said that menstruating girls were not allowed to use the hostel toilets for changing their absorbents. She added,
>
> “Sometimes we used to climb up a hillock just to relieve ourselves or to change the absorbents.”

All the married (for not more than three months) girls from tribal area whom we interviewed were unsure regarding whether their husbands knew about menstruation. None of them had tried to bring this topic up and had only shared their date for periods with their mother-in-law. The already gendered experience of menstruation that girls face within households seemed to become more stressful as girls got married and moved to their in-laws’ households. A 19-year-old married girl from tribal setting shared,

> “Here everything is new… I will have to tell my mother-in-law when I get my periods as she will tell me what to do… here the house is new for me. I don’t know, where to wash my menstrual cloth, where to dry it so that no one sees it. One day I will have to talk to my husband about it because you know…. Married men should know about it…. if the man wants to do something (sexual intercourse), he will still do it even if his wife is menstruating”

Marital status of the adolescent girls also seemed to contribute to the gendered perception of the self and the body among the tribal poor. Although in the pre-survey qualitative phase, we did not come across any girl (all unmarried) mentioning pubic hair removal as the essential step in menstrual hygiene, married girls from the post-survey phase did mention it as a necessary step. At least one married tribal girl confirmed buying a hair removal crème every month for ‘cleaning’ the area, though it was economically taxing. She said she “had to somehow manage money to buy one.”

We shared that the teasing experience narrated by several menstruating girls was mostly denied by boys, as reflected in our results based on both the qualitative and the quantitative pieces. Girl participants in the post-qualitative phase reiterated that the teasing experience was indeed common and affected several girls emotionally. A girl participant from tribal area narrated incident when they witnessed their friend crying as she was teased (laughter upon seeing a stain). A married tribal girl shared a teasing incident faced by her school-friend and wondered whether boys tease because they don’t know that menstrual blood flow is not a bodily fluid that can be controlled by the body (such as urinary control). She wondered whether they laugh because they think that it is girls’ immaturity to have a stain as she “suddenly loses control over her body (sphincters).”

The social worker supported our findings regarding gender being a social determinant of menstruation-related experience. He explained,

> “It [being a boy] makes society adamant about what he can participate in. Being a woman is at least beneficial in the sense that women come together and discuss about menstruation. I find it extremely challenging to gather boys from disadvantaged backgrounds for sex education as their coming together is always politicized. Family resists if we try to get them out of the rigid cage of masculinity.. In this community we also see a close link between menstruation and domestic violence.”

Key informants also pointed out that one needs to examine the complexity in the relationship amongst caste, class, and menstrual experience. The sex-education expert cautioned that while tribal societies are considered egalitarian, one needs to find out if menstruation-related negative beliefs are due to infrequent water supply, a lack of privacy, or other such constraints, rather than social reasons. The social worker explained that class and wealth may not show an incremental relationship with menstruation-related outcomes because stereotypes regarding gender, caste, class, and honour intersect in complex ways.Parents’ lack of consent for inclusion of their school-dropout sons in our interviews also hinted at greater barriers to healthy SRH faced by school-dropout boys.

## Discussion

We found that gender influences adolescents’ menstruation-related experiences, understanding, and beliefs. It unpacks the various layers explaining how menstruation becomes a gendered experience in different social domains for girls and influences their health. The quantitative findings highlight the gender differences in knowledge, beliefs, receipt of information in schools and also in opinion on ‘whether boys should know about menstruation’. Further, the quantitative strand unearthed how gendered experience of menstruation influenced menstrual health-related outcomes.

Thus, our analysis added textured explanations to support the previous findings suggesting that gender is an important determinant of sexual and reproductive health among adolescents; operating through unjust roles, norms, and expectations.

The gender analysis framework guided us in disentangling the complex ways in which being a boy/girl influences menstrual health-related experiences: by (i) governing adolescents’ access to information, ability to possess knowledge, and ability to possess beliefs, (ii) creating gender roles, expectations, and by sustaining stigma, (iii) by negatively impacting ‘girls’ menstrual beliefs and health outcomes.

Our finding that being a boy (than a girl) likely meant not receiving information in schools and not possessing adequate and accurate knowledge, corroborates the findings of previous studies on Indian adolescents’ sexual and reproductive health (SRH). Additionally, the finding that boys were curious to learn about reproductive health (a greater proportion of boys versus girls opined that boys should know about menstruation) was similar to previous studies on Indian boys (S. E. Mahon et al., 2015; Mason et al., 2013). Notably, girls expressed a fear of getting teased by boys if boys got to know about menstruation. Our pre- survey as well as the post-survey qualitative phase reflected that teasing faced by girls could have several layers to it as it could emerge from boys’ awkwardness, their increased sense of differences between a male and a female body, as well as their unawareness regarding the nuanced aspects of menstruation (for instance, that menstrual blood flow cannot be ‘controlled’ by a woman’s body). Our study brings forth that gender inequality was produced and sustained as menstruation was viewed as a biosocial marker to differentiate what girls and boys can/ not do, an important finding corroborating previous research on adolescents (T. Mahon & Fernandes, 2010; Pradhan & Ram, 2010; Santhya & Jejeebhoy, 2014). This analysis also shows that girls felt awkward to interact with-and hid their menstrual status and absorbents from-men, which also echoes the finding by scholars who argue that by concealing their menstrual status, girls learn to un-gender their sexual maturity, while also subscribing to the societal expectations of the gendered power relations that generate stigma for their bodily processes and subordination to men (Caruso et al., 2017; Dube, 1988; Fingerson, 2006; Luke, 1997). Our post-survey qualitative phase unpacked how marital status added more burden to girls’ already gendered menstrual experience as they struggled to fit in with the normative ideas of cleanliness and menstrual hygiene management (for instance, by using hair removal cream for removing their pubic hair), while remaining clueless about how to discuss menstruation with their husbands. The influence of the political economy of the hygiene industry on the ideas of cleanliness was reflected in our data as girls’ perceptions regarding their own body and bodily processes seemed to get affected by the created notions of hygiene (Lahiri-Dutt, 2014).

Our qualitative data also corroborated the multi-dimensional nature of the gendered experience, influenced by different axes of determinants fluidly intersecting in a given situation (Sen & Iyer, 2012). Girls’ ability to push the boundaries in avoiding a gendered experience of menstruation within their communities could be limited due to several social and familial inequalities intersecting with gender inequality. For instance, residing or passing through a neighbourhood perceived as unsafe may limit a menstruating girl’s ability to freely talk to boys. Research on sanitation insecurity in India has revealed that girls from the tribal setting often feel insecure using sanitation facilities outside of homes or to defecate in the open as that increases their risk of interactions with intoxicated men (Caruso et al., 2017; Sahoo et al., 2015). Although tribal societies are largely viewed as egalitarian in nature, it has been found that girls and women find it difficult to transfer any egalitarian views on SRH from one social institution to the other due to the difference in the nature of the gender-related power dynamic across social institutions that lessens their scope to bargain with patriarchal discriminatory practices (Caruso et al., 2017; Maharatna, 2000; Richards et al., 2013). Also, girls and women not speaking to men in the open is often normalized and an attempt to break this norm is found to be challenging by most girls and women, especially from the disadvantaged backgrounds (Scott et al., 2017). A study from North India which tested the participatory approach of Village Health, Sanitation, and Nutrition Committees (VHSNCs) to challenge the gendered social experience faced by women members of the committees, found it to be effective only among the few women who could push the strict gender boundaries (Scott et al., 2017). This was primarily associated with women’s realization of their limited ability to bargain with the patriarchy as they had to return to their homes where gender power relations were likely less negotiable. Such dynamics may have influenced even adolescent girls’ experiences due to- or post- menarche.

Moreover, the process by which gender might intersect with caste and class is not one-dimensional. Scholarship on intersectionality indicates that lower class, caste, and gender might intersect to give rise to higher morbidities among women belonging to poor SES (Scott et al., 2017). On the other hand, literature also suggests that an intersection of higher class and patriarchy might limit a woman’s ability to seek autonomy and agency; as such a woman’s seclusion is largely associated with the honour of the family. These dimensions are perhaps reflected as girls from an urban setting going to private schools whose mothers are moderately educated facing gendered experience of menstruation, and also those belonging to a reserved caste category or the fact that girls in urban private schools whose mothers are moderately educated and belong to open category reported feeling ashamed to buy sanitary napkins from a male shopkeeper.

This suggests that at both the higher and the lower levels of class and caste, women seem to be less valued. While the minute processes through which this complex dynamic becomes a social experience may differ in different social environments, we could find a glimpse of this complexity in our data.

An essential part of the intersectional nature of social identities was showcased by our qualitative data shedding light on the several ways through which being a boy also invited gendered experiences: boys remained deprived of information regarding menstruation and their curiosity remained unaddressed. Thus, a gendered experience is not restricted to girls but boys, too, were adversely affected in several ways by the gendered nature of menstruation-communication. Intersectionality scholars highlight that *gender and health* is not necessarily about *women* and *women’s health* (Hankivsky, 2012).

Underscoring the gendered nature of menstruation, this article shows that the discrimination that girls face affects their menstrual beliefs and health. Our findings shed light on a few mechanisms explaining how gender inequality affects women’s health, links also discussed by several scholars in the past (Chandra-Mouli et al., 2013; Krieger, 2003; Moss, 2002; Sanneving et al., 2013; Tolman et al., 2003). A study from Pondicherry (n=800) showcased how a ‘traditionality index,’ indicating gendered beliefs regarding menstruation, was higher among rural and poor girls and a greater proportion (36%) of ‘very traditional’ girls reported white discharge during menstruation as compared to ‘very modern’ (18.1%) (Narayan et al., 2001). This closely reflects our findings that the experience of gendered menstruation reduces the chance of possessing good menstrual health. However, in this chapter we add further layers as we look at (i) the heterogeneity in gender-based discrimination in different social environments and across socioeconomic settings; (ii) the influence of school-level social determinants in addition to parental socioeconomic characteristics; (iii) the impact of gendered menstruation on possessing positive beliefs; and, (iv) menstrual health status as a measure reflecting many aspects of menstrual health than only white discharge.

Our ability to generalize the findings to a wider population is limited because we used a non-probabilistic sampling design and could not include school drop-outs and adolescents less than 12 years old. However, our sample was adequate to detect socioeconomic patterns in the gendered experience of menstruation, a unique contribution in the Indian context. The cross-sectional nature of this study limits our ability to study the causal pathways. However, as a first attempt at unpacking how gender is a social determinant of menstrual health, this study still adds value. Our integrated theoretical approach does not capture the psychological and developmental aspects of adolescents’ experiences regarding menstruation; however, it does justice to the application of a social epidemiological lens. Knowledge, beliefs, and gendered menstruation index variables were calculated by an additive method, which might not represent the complexities of these constructs. Additionally, variables for parental socioeconomic characteristics were reported by adolescents who might have failed to give adequate and accurate information (10% of wealth data were missing). Our inability to cross-check this information by referring to school-records remains a limitation. Furthermore, when compared with a data representative of Nashik district, our sample lack a representation of the poorest two categories of household wealth because we used a school-based survey which excluded poor adolescents who are most likely school drop-outs. This potentially is a reason for the inconsistent and weak findings pertaining to parents’ socioeconomic characteristics and wealth. Our sample size also prevented us from performing subgroup analyses examining how gender and social inequalities interact. Additionally, due to limited resources, we were unable to gather data on objectively assessed menstrual health. Despite these limitations, we could highlight how gender is a social determinant of menstrual health, how various social factors (mother’s education, setting, wealth) create patterns in the menstrual experience of girls in different social environments (family, school, community) and how this gendered experience affects girls’ health outcomes (beliefs and menstrual health status).

To our knowledge, this is the first mixed-methods study using a gender-sensitive methodology and an integrated theoretical approach that draws attention to how gender and social inequalities intersect to impact girls ‘and boys’ understanding, beliefs and experiences regarding menstruation, a unique social epidemiological approach that questions treating menstruation as a ‘women’s only’ topic. By using a mixed-methods design, we could unearth context-specific nuances of the gendered social experience of menstruation while quantifying the social patterning in this gendered experience and its influence on girls’ menstrual health. The mixing enabled a coherent interpretation of these nuances and patterns. Our finding that the gendered experience of menstruation differs in different social domains highlights how gender permeates structural spaces and environments and acts as an “organizing principle of social life” (Gita et al., 2002; Sims & Butter, 2002).

## Conclusion

Our study showcases how menstrual health, which is largely considered a ‘women’s’ topic, reflects unjust gender and socioeconomic differences in the accessibility of information for both boys and girls at a stage in their lives when they learn to conform to expectations.These unjust differences rooted in the gender inequalities perpetuated in patriarchal societies such as India govern girls’ menstruation by treating it as a shameful experience.

Thus, by recognizing the role of boys and men in creating and sustaining taboos around menstruation, this study underscores the need to engage them in creating menstruation-friendly societies. Although programs exist which target narrowing gender-based discrimination and gender gaps in health, several programs focus on developing skillsets among women, improving their access to financial resources but fewer programs aiming to reduce gender disparities reach marginalized adolescent girls and boys who have specific needs. Although India’s national-level program for adolescents, the *Rashtriya Kishor Swasthya Karyakram*, recommends involving boys in the discussion on menstruation, it is not based on evidence regarding the similarities and differences in girls’ and boys’ menstruation-related understanding (Ministry of Health and Family Welfare, 2017). It is only by recognizing adolescents’ needs specific to their gender and socio-demographic characteristics that patriarchal societies such as India can aim to develop gender-sensitive menstrual health strategies for Indian adolescents.

## Data Availability

Data cannot be shared publicly because of the sensitive nature and the confidentiality issues. Specifically, our participants and their responsible adults in this study did not give consent for their data to be released to anyone other than the research team as this dataset contains sensitive information. In order to collect meaningful data, assurances regarding participant privacy were critical.

